# Prevalence and Predictors of Preterm Delivery among Mothers at the Tamale Teaching Hospital in the Tamale Metropolis of Ghana

**DOI:** 10.1101/2024.06.01.24308321

**Authors:** George Sarpong Agyemang, Jonathan Mawutor Gmanyami, Elvis Angelo Appiah, Samuel Adolf Bosoka, Mawuli Gohoho, James Alorwu, Amos Ziema Sorengmen, Umar Haruna, Michael Yaw Amoakoh, Margaret Kweku

## Abstract

**Background:** Preterm delivery (PTD) remains a global public health challenge. It is estimated that 15 million babies are born prematurely. Developing countries account for 18.3% of PTD. This study assessed the prevalence and risk factors associated with PTD deliveries in the Tamale Metropolis.

**Methods:** A cross-sectional study accessed the birth records and interviewed 318 participants who delivered at the Tamale Teaching Hospital between January to December 2021. Univariate and multivariate regression models predicted the risk factors of PTD.

**Results:** The prevalence of preterm delivery was 18.5%. Women with multiple pregnancies who had a caesarean section and large household size were 4.28, 7.31 and 6.88, times more likely to have preterm delivery [AOR = 4.28 (95% CI: 1.75-24.23)], [AOR = 7.31 (95% CI: 2.59-36.14)] and [AOR = 6.88 (95% CI: 1.56-30.38)] respectively. Women who had normal haemoglobin concentration levels at 36 weeks and those who had pipe-borne water as the source of drinking water were 90% and 78% less likely to have preterm delivery [AOR = 0.10 (95% CI: 0.03-0.33)] and [AOR = 0.22 (95% CI: 0.06-0.78)] respectively.

**Conclusion:** There is a high prevalence of preterm delivery in the Tamale Metropolis. Having multiple pregnancies, caesarean sections and large household size are risk factors of preterm birth. Whilst maintaining a normal haemoglobin at 36 weeks and drinking pipe-borne water are protective factors against preterm delivery in the Tamale metropolis.

## Introduction

Preterm delivery (PTD) is defined as babies born alive before 37 weeks of pregnancy are completed (1). Every year, an estimated 15 million babies are born prematurely. In 2010, 14.9 million babies were born preterm, accounting for 11.1% of all births worldwide (2). These birth rates vary from approximately 5% in various European nations to 18% in other African nations. South Asia and sub-Saharan Africa account for 52% of all live births worldwide, and more than 60% of preterm infants are born (2). Developed countries are also affected by preterm birth; the United States and Austria reported a high prevalence of 12.0% and 9.3%, respectively (2). However, the problem is more prevalent in developing countries than in developed countries where a prevalence of 18.3% has been reported in Kenya (3).

A million children die each year from problems associated with preterm delivery (4). Preterm birth has been reported to be the leading cause of neonatal mortality. Infants born preterm are four times more likely to die during the neonatal period than term infants (2). Infants with PTD have a higher risk of developmental disabilities, including cerebral palsy and retinopathy of prematurity. The consequences of preterm birth may continue into adulthood, increasing the risk of adult-onset chronic conditions such as obesity and diabetes (1). According to the World Health Organization (WHO), the aetiology of PTD remains unclear (1). However, some researchers argue that morbidities and infections such as malaria, hypertension, syphilis, and HIV can cause preterm delivery (5,6). Others argue that PTD is caused by multiple aetiologies such as individual and environmental factors (7,8). Some studies have reported that factors such as age, socioeconomic status, gestational diabetes, preeclampsia, and foetal distress influence PTD (3).

PTD is an important public health concern in Ghana. Although interventions to reduce infant mortality have been instituted, this problem persists. This is evident in studies conducted by (9) in Ho and Accra by (10), who reported a prevalence of 14.1% and 37.3%, respectively. According to the 2014 GDHS (11), the Northern Region is reportedly one of the regions with the worst health indicators for newborns and under-five mortality rates. Preterm delivery is the leading cause of death in children under the age of 5 years (12). Thus, complications associated with preterm birth could be the reason for the poor neonatal indicators in the Northern Region. Based on the different prevalence of preterm births reported by previous studies and the dearth of research on this subject matter in the region, it is vital to study the factors that influence this outcome.

Identifying the risk factors for preterm delivery is crucial in providing appropriate care and attention to pregnant women at risk. Unfortunately, little is known about these factors in the Tamale Metropolis, which can jeopardise the future of newborns. By identifying these factors, we can significantly reduce the incidence of preterm babies, associated costs, and the burden on the healthcare system. This will, in turn, lower the rates of neonatal and childhood morbidity and mortality throughout the region and country. Achieving this goal will aid in meeting Sustainable Development Goal 3’s target of reaching a neonatal mortality rate of 12 per 1,000 live births by 2030. Therefore, this study assessed the prevalence and factors associated with preterm delivery in the Tamale Metropolis of Ghana.

## Materials and Methods

### Study Setting

The study was conducted at the Tamale Teaching Hospital (TTH) in the Northern Region of Ghana from 1st February 2022 to 31 May 2022. TTH is the primary referral hospital in the five Northern regions of the country and was established in 1974. The hospital has a bed capacity of around 800 and provides specialist services in various fields, including obstetrics and gynaecology, surgery, orthopaedics and trauma, internal medicine, child health, pathology, ear, nose and throat, eye unit, endoscopy, neurosurgery, anaesthesia and intensive care unit, psychiatry, dentistry, pharmacy, laboratory, and outpatient services.

The Tamale Metropolis, where the hospital is located, is one of the 16 districts in the Northern Region and has a population of 374,744. The population is almost equally divided between males (49.4%) and females (50.6%). The Tamale Metropolis has 21 health facilities, including 13 Community-based Health Planning and Services (CHPS) compounds, 4 health centres, 1 Christian Health Association of Ghana (CHAG) hospital, 2 metropolitan hospitals, and 1 teaching hospital.

### Study design

This was a cross-sectional study involving women who delivered in the TTH from January to December 2021. The study was carried out from 1st February 2022 to 31 May, 2022 as the start and end of the recruitment period for this study respectively and examined the birth records of all mothers who delivered live babies at the TTH. Data was extracted from the delivery and ANC registers of mothers using a checklist. In addition, using a semi-structured questionnaire, mothers were followed up and interviewed on information that could not be found or missed in their birth records.

### Study population

The study population included all mothers who gave birth to live-born infants at Tamale Teaching Hospital between January and December 2021.

### Inclusion and Exclusion criteria

In this study, all live births during the specified period were included, except for stillbirths, babies with congenital abnormalities (identified at birth), and birth records with incomplete information. Babies with congenital abnormalities were excluded due to their different risk factors and causes for preterm delivery. Stillbirths were also excluded due to missing gestational age data. Home deliveries were not included because they were not supervised by health professionals, therefore data on maternal and child health variables, including gestation, were not available.

### Sample size calculation

The sample size was calculated using a single population proportion formula by considering the confidence level (95%), the margin of error = 5%, and the 18.3% prevalence obtained from the previous study conducted in Nairobi, Ethiopia (3).

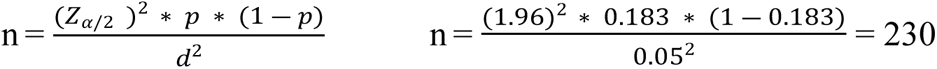

After adding a non-response rate of 10%, the final sample size was 253. However, a sample of 318 was used for the study.

### Sampling Procedure

Systematic random sampling was used to select participants from a list of all eligible mothers. Mothers who delivered live-born babies formed the sampling frame of 1323 for the study. The population size in the sample frame was divided by the sample size to determine the interval for selecting samples for the study. Thus, at every 15^th^ interval in the sampling frame, the participants who met the inclusion criteria were selected for the study. This was done until a total sample size of 318 was achieved.

### Data collection procedure

A semi-structured questionnaire and a checklist which was developed into an extraction sheet which was used to extract data from the ANC register at the ANC clinic and the delivery register at the labour ward. The questionnaire was pretested at the Seventh Day Adventist Hospital in Tamale and was used to interview mothers at home. The data collected included information such as the gestational age at delivery, socio-demographic details such as age, location, marital status, occupation, and contact information, and obstetric characteristics such as parity, gravidity, delivery type, etc. Respondents who met the study’s inclusion criteria and were selected for the study were traced to their homes, and a semi-structured questionnaire was administered to collect additional data missed in the ANC booklet or the labour register. Trained data collectors, proficient in both Dagbani and English, were responsible for administering the questionnaire to the respondents. Eight data collectors were used for this process.

### Definition of key variables

Term delivery: Term delivery is delivery after 37 weeks gestation.

Preterm delivery: Preterm delivery is delivery below 37 weeks of gestation.

Gestational age: Gestation is the period between conception and birth.

Multiple pregnancy: Multiple pregnancy is defined as pregnancy with more than one foetus.

### Study variables

#### Outcome variable

The dependent variable was preterm delivery. Gestational age was dichotomised into 0 “carried to term” when the gestational age at delivery was ≥37 weeks and “preterm delivery” when the gestational age at delivery was <37 weeks. Gestational age was determined at birth from the birth records documented by midwives. Birth records that documented ultrasound to determine gestational age were included.

#### Explanatory variables

Explanatory variables included maternal sociodemographic information such as age, educational level, ethnicity, marital status, religion, place of residence, household number and socioeconomic status. Obstetric characteristics included such as gravidity, type of birth, gravidity, parity (0 =nulliparous, ≥1 = multiparous), pregnancy interval, planned conception and mode of delivery, previous history of abortion, previous history of stillbirth, previous history of complications, family history of hypertension, and LLINS use. Additionally maternal conditions such as hepatitis B, hypertension (Absent=120/80 mmHg and below, Present= above 140/90 mmHg), haemoglobin level (normal ≥ 11.0 g/dl, Anaemia <11.0 g/dl), sickling status, HIV and malaria. Data on maternal conditions were extracted based on routine testing of pregnant women during antenatal care visits. In addition, environmental and lifestyle factors, such as alcohol intake, source of water, kitchen type, month of pregnancy, and cigarette smoking, were obtained through interviews with respondents.

### Data Management and Analysis

The Kobo Collect app was used for data entry, and the data was then exported to STATA version 17 for analysis. Descriptive statistics were used to calculate frequencies, and percentages were reported for categorical variables. For continuous variables, means and standard deviations were determined. To determine the mother’s economic status, various socioeconomic indicator variables were considered Principal component analysis (PCA) was used on the relevant socioeconomic indicator variables that contributed to the combined socioeconomic status score factor by more than 10%. The socio-economic status of a household was described by the factor of the PCA with the highest eigenvalue, and the respective factor scores were divided into terciles and used in the regression analysis. The lowest 33% of households according to the economic status variable were classified as low, and the highest 33% as having high socioeconomic status.

Univariate logistic regression analysis was used to identify associations between variables and PTD (Model I), followed by a stepwise multivariate logistic regression model (Model II) that considered variables that were significant in univariate logistic regression. The goodness of fit of Model II was examined using the likelihood ratio test by comparing the likelihood of data under the full and alternative models. If the overall model recorded a p-value less than 0.05, the model was considered good. The variable inflation factor (VIF) was used to cater for multicollinearity, and explanatory variables with VIF values exceeding 5 were excluded from Model II. Adjusted odds ratios and confidence intervals were computed with p values <0.05 and were considered statistically significant in Model II.

### Ethical consideration

Ethical approval for this study was obtained from the Department of Research and Development, Tamale Teaching Hospital. The ethical clearance number for this study was TTH/R&D/SR/152. Written informed consent was obtained from the respondents before conducting the interviews. Study participants who were less than 18 years, consent was obtain from their guardians or parents. The data obtained from the study was kept confidential, and personal information was anonymised during the data collection, analysis, and dissemination of findings. Identification codes were used for data management, storage, analysis, and reporting. A file containing information about the participants was stored in a cabinet and kept under lock. The file was accessible only to the principal investigator.

## Results

### Socio-demographic characteristics of respondents

Table 1 shows the socio-demographic characteristics of the participants. Three hundred and eighteen women were involved in this study, with a mean age of 28.0 ± 5.2. The majority of participants were aged below 30. A few women (36.2%) had no formal education. The majority of the participants 227 (71.4%) were married and 275 (86.5%) were Mole-Dagombas. Similarly, the majority 286 (89.8%) were Muslims. Most of the participants 211 (66.4%) lived in urban areas within the Tamale Metropolis, with the majority living in households with 1-6 inhabitants. The majority of participants 197 (61.9%) belonged to the low socioeconomic bracket.

**Table 1:** Socio-demographic characteristics of participants.

| Characteristics | Frequency (N=318) | Percentage (%) |
| --- | --- | --- |
| **Age at delivery** |  |  |
| <30 | 226 | 71.1 |
| 30 above | 92 | 28.9 |
| **Educational level** |  |  |
| No Formal Education | 115 | 36.2 |
| Formal education | 203 | 63.8 |
| **Marital status** |  |  |
| Single | 91 | 28.6 |
| Married | 227 | 71.4 |
| **Ethnicity** |  |  |
| Others | 43 | 13.5 |
| Mole-Dagbani | 275 | 86.5 |
| **Religion** |  |  |
| Islam | 286 | 89.9 |
| Christianity | 32 | 10.1 |
| **Place of residence** |  |  |
| Urban | 211 | 66.4 |
| Rural | 107 | 33.6 |
| **Household number** |  |  |
| 1-6 | 182 | 57.2 |
| 7 above | 136 | 42.8 |
| **Socioeconomic status** |  |  |
| Low | 197 | 61.9 |
| High | 121 | 38.1 |

### Maternal Obstetric and Medical Characteristics of Participants

Table 2 shows the obstetric and medical characteristics of the participants. The majority of participants 305 (95.9%) had singleton deliveries. More than half of the participants 202 (63.5%) had 1-2 gravida, with the majority 213 (67.0%) being multiparous. Similarly, the majority of participants (203) had two deliveries in the last five years. However, the majority of the participants 205 (65.5%) planned their conception. Only a few participants 28 (8.8%) delivered through caesarean delivery. Few women had a history of abortion (n = 26, 8.2%), stillbirth (n = 2, 0.6%), and complications (n = 10, 3.1%). Similarly, only a few participants had a family history of hypertension 17 (5.4%). In addition, the majority of the women (241 [75.8 %]) slept in LLINs during pregnancy. With maternal conditions, only a few participants had hepatitis B (n = 8, 2.5%), hypertension during pregnancy (n = 10, 3.1%), anaemia at 36 weeks (n = 132, 41.5%), sickle cell (n = 7, 2.2%), HIV positivity (n = 10, 3.1%), and malaria during their pregnancy (n = 23, 7.2%).

**Table 2:** Obstetric and conditions of participants.

| Characteristics | Frequency (N=318) | Percentage (%) |
| --- | --- | --- |
| **Type of birth** |  |  |
| Single | 305 | 95.9 |
| Twin | 13 | 4.1 |
| **Gravida** |  |  |
| 1-2 | 202 | 63.5 |
| 3+ | 116 | 36.5 |
| **Parity** |  |  |
| Nulliparity | 105 | 33.0 |
| Multiparity | 213 | 67.0 |
| **Pregnancies in the last 5 years** |  |  |
| 1 | 115 | 36.2 |
| 2 above | 203 | 63.8 |
| **Plan conception** |  |  |
| No | 113 | 35.5 |
| Yes | 205 | 65.5 |
| **Mode of delivery** |  |  |
| Spontaneous vaginal delivery | 290 | 91.2 |
| Caesarean section | 28 | 8.8 |
| **Previous history of abortion** |  |  |
| No | 292 | 91.8 |
| Yes | 26 | 8.2 |
| **Previous history of stillbirth** |  |  |
| No | 316 | 99.4 |
| Yes | 2 | 0.6 |
| **Previous history of complications** |  |  |
| Absent | 308 | 96.9 |
| Present | 10 | 3.1 |
| **Family history of hypertension** |  |  |
| No | 301 | 94.7 |
| Yes | 17 | 5.4 |
| **LLINs usage** |  |  |
| No | 77 | 24.2 |
| Yes | 241 | 75.8 |
| **Hepatitis status** |  |  |
| Negative | 310 | 97.5 |
| Positive | 8 | 2.5 |
| **Hypertension during pregnancy** |  |  |
| Absent | 308 | 96.9 |
| Present | 10 | 3.1 |
| **Haemoglobin level at 36 weeks** |  |  |
| Anaemic | 132 | 41.5 |
| Normal | 186 | 58.5 |
| **Sickling status** |  |  |
| Negative | 311 | 97.8 |
| Positive | 7 | 2.2 |
| **HIV status** |  |  |
| Negative | 308 | 96.9 |
| Positive | 10 | 3.1 |
| **Malaria during pregnancy** |  |  |
| Absent | 295 | 92.8 |
| Present | 23 | 7.2 |

### Environmental and Lifestyle characteristics of participants

Table 3 shows the environmental and lifestyle characteristics of participants. Of the respondents, 9 (2.8%) consumed alcohol, with the majority consuming water from pipe source 266 (83.6%). The majority of participants (243 [76.4 %]) used an outdoor kitchen. The majority lived in compound houses 291 (91.5%), while a few of the participants (74 [23.3 %]) used a water closet. The majority of respondents 243 (76.4%) became pregnant during the rainy season. Only 3 (0.3%) women smoked cigarettes during pregnancy.

**Table 3:**
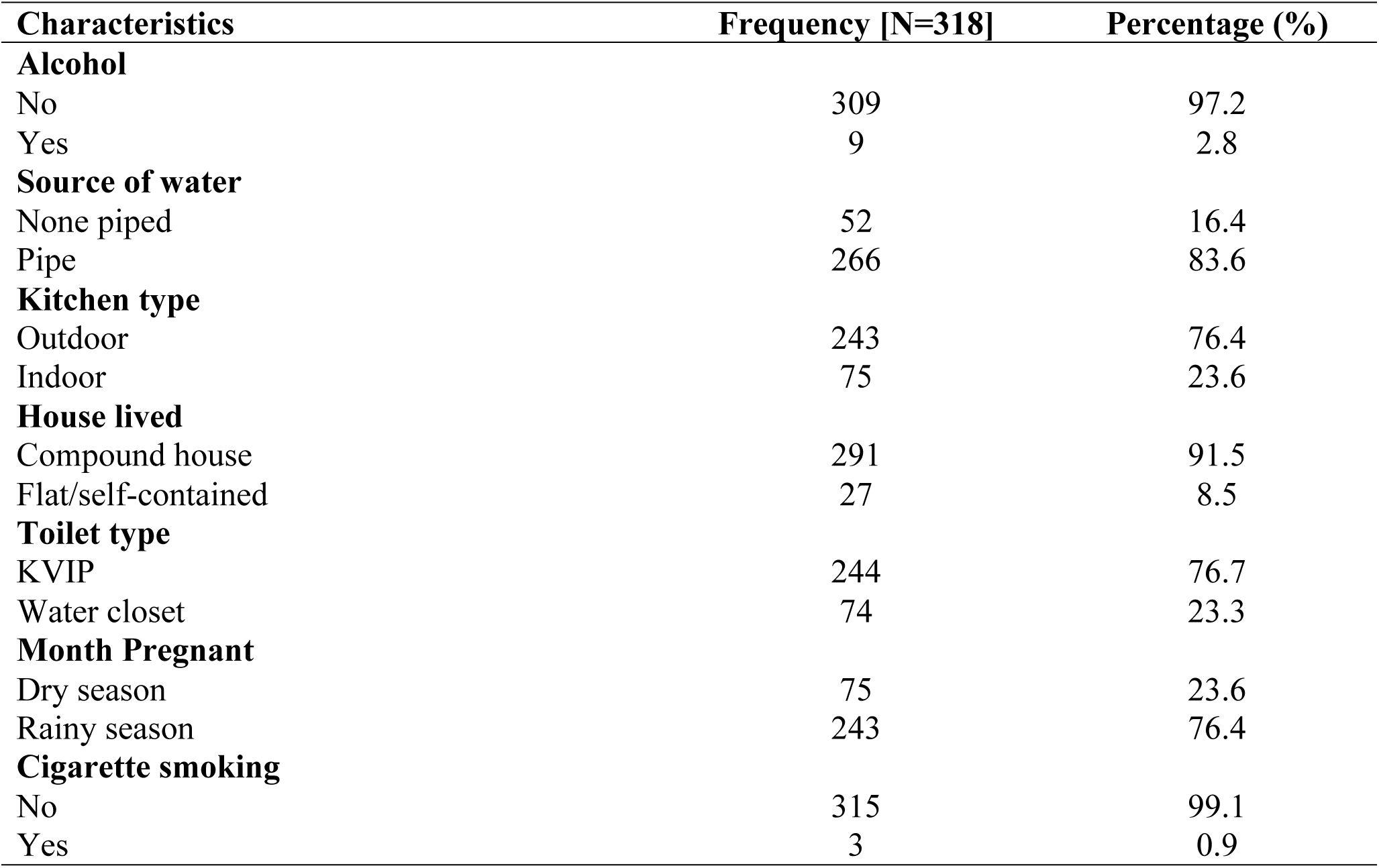
Environmental and lifestyle characteristics of participants.

### Prevalence of preterm birth

Figure 2 shows the prevalence of preterm delivery within the Tamale Metropolis. From the study, an 18.5% prevalence of preterm delivery was reported.

**Figure 2:**
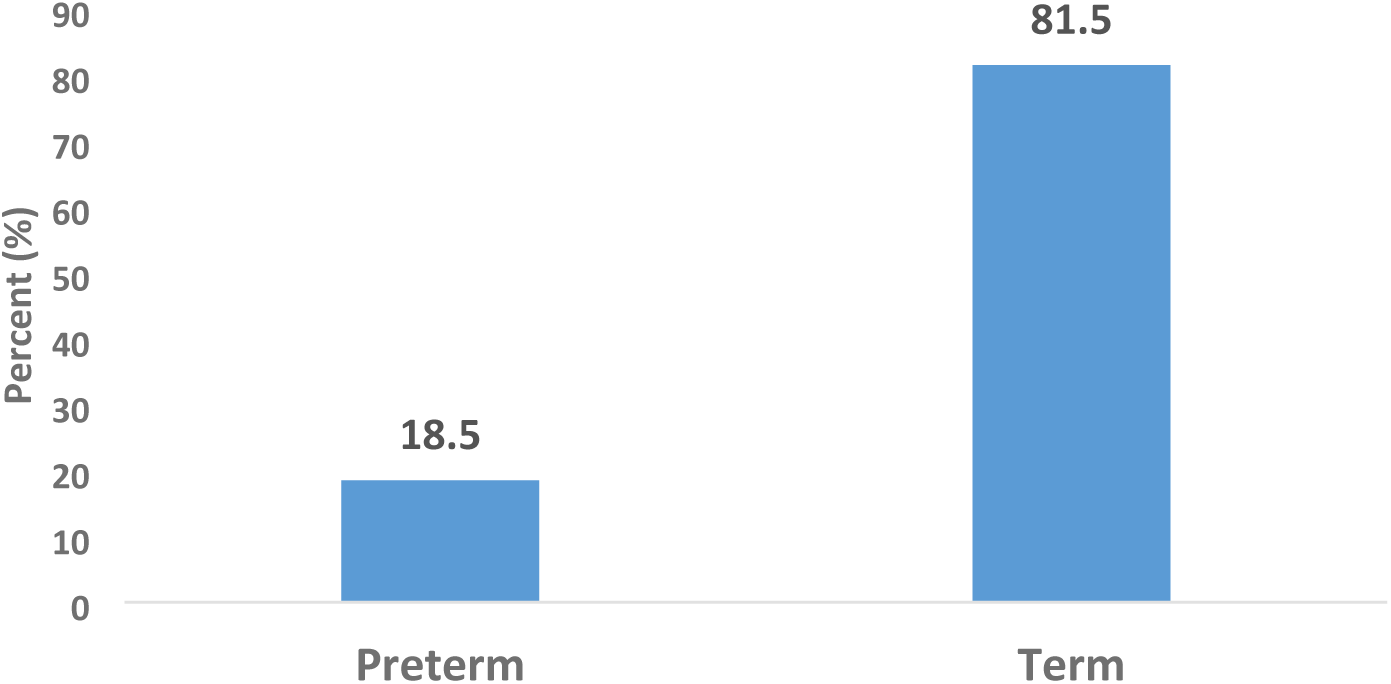

Proportion of mothers with preterm delivery in the Tamale Teaching Hospital

### Factors associated with preterm birth

Table 4 shows that household number (family size), type of birth, mode of delivery, haemoglobin level at 36 weeks, and source of water were factors significantly associated with preterm births. Women who lived in households with 7 or more people were 6 times more likely to have preterm birth as compared to women who lived in households with less than 7 people. And the difference is statistically significant [AOR=6.88 (95% CI: 1.56-30.38), p= 0.011]. Similarly, women with twin delivery (multiple pregnancies) were 4.28 times more likely to have preterm birth as compared to those women with single births and the difference was statistically significant [AOR=4.28 (95% CI: 1.75-24.23), p= 0.025]. Also, women who went through caesarean section were 7.31 times more likely to deliver preterm babies as compared to those who had spontaneous vaginal delivery and the difference was statistically significant [AOR=7.31 (95% CI: 2.59-36.14), p= 0.008].

**Table 4:** Factors associated with preterm delivery in the Tamale Metropolis.

| Variable | Gestation Age |  | COR [95% CI], P-value | AOR [95, P-value |
| --- | --- | --- | --- | --- |
|  | Term<br>(N=259)<br>n (%) | Preterm<br>(N=59)<br>n (%) |  |  |
| **Household size** |  |  |  |  |
| 1-6 (small family size) | 157 (60.6) | 25 (42.4) | Ref. | Ref. |
| 7+ (Large family size) | 102 (39.4) | 34 (57.6) | **2.88 (1.33-6.22), 0.007** | **6.88 (1.56-30.38), 0.011** |
| Single | 254 (98.1) | 51 (86.4) | Ref. | Ref. |
| Multiple | 5 (1.9) | 8 (13.6) | **6.54 (1.80-17.26), 0.012** | **4.28 (1.75-24.23), 0.025** |
| Spontaneous vaginal delivery | 247 (95.4) | 43 (72.9) | Ref. | Ref. |
| Caesarean section | 12 (4.6) | 16 (27.1) | **12.39 (2.80-44.73), 0.001** | **7.31 (2.59-36.14), 0.008** |
| Positive | 5 (1.9) | 3 (5.1) |  |  |
| **Hypertension at ANC** |  |  |  |  |
| Absent | 254 (98.1) | 54 (91.5) | Ref. |  |
| Present | 5 (1.9) | 5 (8.5) | 4.32 (0.82-22.81), 0.084 |  |
| **Haemoglobin level at 36 weeks** |  |  |  |  |
| Anaemic | 84 (32.4) | 48 (81.4) | Ref. | Ref. |
| Normal | 175 (67.6) | 11 (18.6) | **0.09 (0.04-0.22), <0.001** | **0.10 (0.03-0.33), <0.001** |
| **Source of water** |  |  |  |  |
| None piped | 34 (13.2) | 17 (29.3) | Ref. | Ref. |
| Pipe | 224 (86.8) | 41 (70.7) | **0.22 (0.09-0.54), 0.001** | **0.22 (0.06-0.78), 0.019** |
| **Kitchen type** |  |  |  |  |
| Outdoor | 192 (74.1) | 51 (86.4) | Ref. |  |
| Indoor | 67 (25.9) | 8 (13.6) | 0.67 (0.27-1.66), 0.389 |  |
| **Toilet type** |  |  |  |  |
| KVIP | 193 (74.5) | 51 (86.4) | Ref. |  |
| Water closet | 66 (25.5) | 8 (13.6) | 0.62 (0.26-1.46), 0.280 |  |

However, women who had a normal haemoglobin level at 36 weeks were 91% less likely to deliver preterm birth as compared to women who were anaemic at 36 weeks and the difference was statistically significant [AOR=0.10 (95% CI: 0.03-0.33), p= <0.001]. Also, women who consumed pipe-borne water during their pregnancy were 78% less likely to have preterm births as compared to those who consumed none-pipe water and the difference is statistically significant [AOR=0.22 (95% CI: 0.09-0.54), p= 0.001].

## Discussion

Our study found a prevalence of preterm delivery of 18.5% among women in the Tamale Metropolis. This is similar to studies in the Volta Region at the Ho Teaching Hospital by (9), which reported a prevalence of 14.1%, notably higher than rates reported in other regions such as the Komfo Anokye Teaching Hospital, which reported a prevalence of 37.3% (10). The similarity in the prevalence of preterm delivery in the Tamale Teaching Hospital could be attributed to the referral nature of the study setting. Despite several interventions to prevent preterm delivery, it is clear the problem persists at the Tamale Teaching Hospital (6). This has the propensity to affect the achievement of SDG goal 3. Women with a high risk of medical and obstetric complications are referred to have advanced management and care in tertiary hospitals in developing countries. Thus, tertiary hospitals must be provided with the needed infrastructure and logistics to be able to manage these complications and provide the needed care and management.

In this study, women with an increasing household number, multiple pregnancies, caesarean section, normal haemoglobin concentration level at 36 weeks, and those whose source of drinking is pipe-borne water were the significant predictors of PTD. Additionally, household number was significantly associated with preterm. Pregnant women who belonged to households of more than seven were 6 times more likely to deliver preterm babies compared to pregnant women who belonged to households of less than seven. This is inconsistent with previous studies conducted in Ethiopia, which found no association between household number and preterm delivery (13–15). The plausible reason for the difference could be because of the differences in the study population, the socio-cultural demographic, and a large household or family size that can have an impact on household food security. Household food insecurity has been linked with household nutrition, which could impact birth weight. Low Birth Weight is significantly associated with preterm delivery (16).

In this current study, the type of birth of the mother was significantly associated with preterm delivery. Women with multiple gestations had higher odds of delivering preterm babies compared with mothers with a single delivery. This is similar to a study conducted in China where the odds of having preterm delivery were 3.3% higher among women with twin births compared with women of single birth (17). The plausible reason for this finding could be that mothers with a twin pregnancy are more likely to suffer pregnancy complications, such as anaemia, hypertensive disorders, gestational diabetes mellitus, and postpartum haemorrhage than those with singleton gestation (18). Also, compared with singleton gestation, twin gestation is associated with a significantly increased risk of perinatal maternal and fetal morbidity and mortality (17).

In this present study, the mode of delivery was significantly associated with preterm delivery. The odds of preterm delivery were higher among women with Caesarean Section compared with women with spontaneous vaginal delivery. This is consistent with a similar study conducted in Ho, Ghana, where they reported that CS increased the odds of preterm delivery by 2 folds compared with a mother who had a spontaneous vaginal delivery (9). A similar study conducted in Spain found that compared to women with spontaneous delivery, women with CS were more likely to have PTD (19). This could be attributed to the abuse of planned CS. This phenomenon has been documented in an earlier study in Brazil, where it was reported that CS was wrongfully associated with preterm delivery, particularly among private hospitals (20). In that regard, there is a need to adhere to WHO’s recommendations that CS birth should not be planned before 39 completed weeks of gestation unless it is medically indicated for the benefit of either the foetus, mother, or both (9).

Haemoglobin level at 36 weeks was significantly associated with preterm delivery. In this study, women with a normal haemoglobin level at 36 weeks were 90% less likely to have PTD. This is consistent with a similar study conducted in Ghana by conducted (9). Their study reported that women with normal haemoglobin were less likely to have PTD compared with women who were anaemic. Elsewhere in China, a study reported that mothers who had a high haemoglobin level during pregnancy had 91% decreased odds of PTD compared with mothers who were anaemic (21). Other studies have also confirmed that anaemia is an independent risk factor for preterm birth, although studies on Hb levels and the risk of preterm birth are sparse. However, an international multicenter cross-sectional study of 5690 singleton and nulliparous pregnancies in four high-income countries, that is, New Zealand, Australia, England, and Ireland, indicated that there was no statistically significant effect of anaemia on the risk of preterm birth (22). The difference in population characteristics of the study participants in the different studies may be the main reason for these inconsistent results.

In this study, the source of drinking water was significantly associated with preterm delivery. The odds of preterm delivery were significantly decreased among women who consumed pipe drinking water compared to women who consumed non-pipe water. Thus, women who drank pipe drinking water were 78% less likely to have PTD compared with women who consumed non-pipe water. In the Tamale Metropolis, drinking water is commonly treated with chlorine to kill bacteria and reduce infections. This source of drinking water is therefore safe from contamination when compared to water from non-pipe sources. In the Tamale metropolis, due to water scarcity in some parts of the metropolis, some women resort to drinking water from dams and wells. These water sources expose them to infections and diseases such as cholera, typhoid fever, and dysentery. Water contaminants in pregnant women can be dangerous as they expose both the mother and foetus to risk. The presence of microorganisms, including bacteria, viruses, parasites, chemicals, and radioactive substances, in drinking water can cause serious complications in pregnant women, leading to preterm delivery (22). This is consistent with a study conducted in California. The study found that higher concentrations of nitrate in drinking water during pregnancy were associated with an increased risk of spontaneous preterm birth (22).

## Conclusion

This study has shown that living in a large household, having multiple pregnancies, and using caesarean as a mode of delivery are contributory factors that expose the woman to preterm birth. However, having normal haemoglobin concentration levels at 36 weeks, and drinking pipe-borne water are contributory factors that protect the woman against preterm birth.

## Limitation

The study could not access the nutritional intake of mothers during their pregnancy as it has been proven to influence the outcome of birth.

## Data Availability

All relevant data are within the manuscript and its Supporting Information files.

## Acknowledgements

We are grateful to the women who took part in this study. The authors are also thankful to the Research Department, at Tamale Teaching Hospital.

## Authors’ contribution

GSA: conceptualisation, statistical analyses, interpreting the data, writing the original draft, final review, and editing. JMG: review and editing. EAA: conceptualisation, review, and editing, SAB: review and editing, MG: review and editing, JA: review and editing, AZS: review and editing, UH: review and editing, MYA: review and editing, MK: conceptualisation, review, and editing. The authors read and approved the final manuscript.

## Data availability

The datasets generated and analyzed during the current study are not publicly available due to limitations of ethical approval involving patient data and anonymity. Still, they are available from the corresponding author at a reasonable request.

## Ethical approval and consent for participation

Ethical approval for this study was obtained from the Department of Research and Development, Tamale Teaching Hospital. The ethical clearance number for this study was TTH/R&D/SR/152. Written Informed consent to participate in the study was obtained from all the participants before enrolment.

## Consent for publication

Not applicable

